# Lower estimated glomerular filtration rate relates to cognitive impairment and brain alterations

**DOI:** 10.1101/2024.09.10.24313312

**Authors:** Shady Rahayel, Rémi Goupil, Dominique Suzanne Genest, Florence Lamarche, Mohsen Agharazii, Violette Ayral, Christina Tremblay, François Madore

**Affiliations:** Department of Medicine, University of Montreal, Montreal, Quebec, Canada, 2900 Edouard Montpetit Blvd, H3T 1J4; Centre for Advanced Research in Sleep Medicine, Hôpital du Sacré-Cœur de Montréal, Montreal, Quebec, Canada, 5400 West Gouin Blvd, H4J 1C5; Division of Nephrology, Hôpital du Sacré-Coeur de Montréal, Montreal, 5400 West Gouin Blvd, H4J 1C5; Divison of Nephrology, Department of Medicine, Université Laval, Quebec City, Quebec, Canada, 2705 Laurier Blvd, G1V 4G2; Centre de Recherche du CHU de Québec-Université Laval, Quebec City, Quebec, Canada, 2705 Laurier Blvd, G1V 4G2; Department of Neuroscience, University of Montreal, Montreal, Quebec, Canada, 2900 Edouard Montpetit Blvd, H3T 1J4; The Neuro (Montreal General Hospital-Institute), McGill University, Montreal, Quebec, Canada, 3801 University St, H3A 2B4

**Keywords:** Chronic kidney disease, glomerular filtration rate, cognition, brain, atrophy, MRI

## Abstract

**Introduction:** Chronic kidney disease is associated with cognitive decline and changes in brain structure. However, their associations remain unclear, particularly the selective vulnerability characteristics that make some brain regions more vulnerable.

**Methods:** We investigated the association between eGFR and cognitive function in 15,897 individuals from the CARTaGENE cohort. We performed vertex-based MRI analyses between eGFR and cortical thickness in the 1,397 participants who underwent brain MRI after six years. Imaging transcriptomics was used to characterize the gene expression and neurodegenerative features associated with this association.

**Results:** Lower eGFR correlated with reduced cognitive performance and brain structure. Brain regions associated with eGFR were enriched for mitochondrial and inflammatory-related genes. These associations occurred independently from age, sex, education, body mass index, Framingham risk score, and white matter lesion volume.

**Discussion:** This study highlights the link between reduced eGFR, cognitive impairment, and brain structure, revealing some of the kidney-brain axis mechanisms.

## 1. INTRODUCTION

Chronic kidney disease (CKD) is characterized by the gradual loss of kidney function, as indicated by lower estimated glomerular filtration rates (eGFR) and/or albuminuria.^1^ It affects 10% of the global population^2^ and is increasingly recognized as a major risk factor for cognitive decline and neurodegenerative conditions such as Alzheimer’s and Parkinson’s diseases.^3–8^ This suggests that CKD has systemic effects that may affect the brain, referred to as the kidney-brain axis. However, the mechanisms associating CKD with cognition and neurodegeneration remain unclear.

Previous studies demonstrated that lower eGFR is associated with changes in brain structure,^9^ manifesting as alterations in white matter tissue, connectivity, and brain atrophy. While the relationship between lower eGFR and decreased white matter integrity has been well-documented,^10, 11^ the relationship with brain atrophy is less clear,^12–15^ especially regarding why certain brain regions are more susceptible to the effects of lower eGFR. Understanding the local features of brain regions more susceptible to develop atrophy is important, as brain atrophy is a hallmark of neurodegenerative diseases affecting cognition.^16^ In degenerative neurocognitive disorders, brain atrophy is not random and has been shown to follow specific constraints dictated by the brain’s regional cellular characteristics, notably through gene expression, and connectivity architecture.^17, 18^ Understanding the features that render some brain regions more vulnerable to atrophy in association with lower eGFR may identify pathways to target for preventing CKD-related neurodegeneration and imaging markers to identify patients at risk of cognitive decline.

In this study, we investigated the relationship between lower eGFR, cognition, and structural brain changes in a population-based longitudinal cohort of 20,004 individuals monitored over six years. We examined the gene expression features of the brain regions associated with lower eGFR levels and assessed the spatial alignment between the eGFR-atrophy brain pattern and the atrophy patterns associated with other neurodegenerative conditions. We hypothesized that reduced eGFR would be associated with lower cognitive performance and brain atrophy and that the spatial pattern of atrophy would be associated with distinct gene expression and neurodegenerative features.

## 2. METHODS

### 2.1. CARTaGENE cohort

The CARTaGENE cohort is a population-based study designed to investigate chronic disease determinants.^19^ This cohort includes over 19,996 individuals aged 40 to 69 years, selected from Quebec’s urban population between 2009 and 2010. At baseline, participants underwent sociodemographic and lifestyle questionnaires, anthropometric measurements, and blood sample collection. After six years, some participants were invited to undergo imaging studies. The current study focused on participants who had complete clinical data from the baseline and underwent a brain MRI scan at follow-up.

### 2.2. Clinical variables

At baseline, standardized questionnaires were used to obtain demographic and medical information. Physical measurements were used to calculate the body mass index and blood samples were collected for biochemical analysis (including glucose, hemoglobin

A1c, and lipid panel) and eGFR quantification, amongst others. The eGFR was computed using the CKD Epidemiology Collaboration formula.^20^ The diagnosis of diabetes was based on a glycosylated hemoglobin A1c level of ≥6.5%, a fasting glucose level of ≥7.0 mmol/L or the use of hypoglycemic medications. The cardiovascular disease risk was assessed using the composite Framingham risk score, which estimates the ten-year risk of cardiovascular disease outcomes by incorporating age, sex, diabetes status, total cholesterol, high-density lipoprotein cholesterol, smoking status, systolic blood pressure, and body mass index.^21^

### 2.3. Cognitive assessment

At baseline, reaction time, paired associates learning, and reasoning tasks were administered through a computerized touch-screen interface. In the reaction time task, participants were required to quickly press a button when a symbol on the screen matched a previously displayed symbol. In the paired associates learning task, participants had to remember and recall the positions of seven targets sequentially presented on the screen. Upon their disappearance, participants indicated their positions. In the reasoning task, participants attempted to solve a series of 12 verbal and numeric reasoning problems within a two-minute time frame.

### 2.4. MRI acquisition and processing

FreeSurfer 7.2.0 was used to process the T1-weighted brain MRI scans to generate whole-brain vertex-based maps of cortical thickness.^22, 23^ Cortical thickness was measured as the distance from the gray/white boundary to the gray/cerebrospinal fluid boundary at each vertex. This cortical thickness approach has been validated against histological analysis and manual measurements,^24, 25^ with consistent reliability found across various scanner types and field strengths.^26, 27^ A 15-mm full-width at half-maximum Gaussian smoothing kernel was applied, and cortical thickness was measured at each vertex to investigate the associations between eGFR and brain morphology across the cortical sheet. The processing also quantified areas of white matter hypointensities using spatial intensity gradients across tissue classes,^28, 29^ an approach that correlates with volumes derived from fluid-attenuated inversion recovery images^30^ and used to account for white matter lesion volume.

### 2.5. Imaging transcriptomics

#### 2.5.1. Regional gene brain expression

To investigate the genetic features of the regions where lower eGFR related to cortical thickness, the cortical maps were parcellated using the Desikan-Killiany atlas to quantify the average thickness across brain regions.^28^ The tool abagen^31^ was next used to extract the brain maps of gene expression of over 20,000 genes, which were quantified across 3,702 tissue samples obtained from six healthy post-mortem brains.^32^ Gene expression values were then normalized across tissue samples. Samples assigned to the same brain region were averaged separately for each donor and averaged across all donors, resulting in a matrix of regional gene expression in the brain. As gene expression data for the right hemisphere was lacking in 4 of the 6 post-mortem brains, the gene expression matrix from the left hemisphere was mirrored onto the right hemisphere and used for the subsequent partial least squares regressions analyses.

#### 2.5.2. Partial least squares regression

Partial least squares regression was used to identify the gene expression components associated with the regional brain patterns representing the association between eGFR and cortical thickness.^33^ This method decomposes two matrices, **X** and **Y**, to derive components from **X** (i.e., the 15,633 genes across the 68 brain regions) that explain the maximal covariance with **Y** (i.e., the 68 regional *t*-values of the eGFR-morphology association).^33^ These *t*-values were derived from linear regression models that accounted for age, sex, body mass index, Framingham risk score, and white matter lesion volume. The components’ significance was tested against the variance explained by 10,000 null models that preserved spatial autocorrelation between brain regions.^34^ A bootstrapping resampling technique with 5,000 repetitions was next applied to identify the genes contributing most to each component. This created a null distribution that enabled the estimation of standard errors for each gene expression weight, with ratios determined by dividing the gene expression weight by its bootstrap-derived standard error.^35^ This ranked list of genes was then used in the functional enrichment analyses.

#### 2.5.3. Functional enrichment analyses

Functional enrichment analyses were used to reveal the biological processes and cellular components enriched in the eGFR-thickness gene expression components. WebGestalt and the Gene Ontology knowledge base^36^ were used to assess whether genes located at the extreme ends of the ranked list were overrepresented in certain biological processes and cellular components beyond what would be expected by chance.^37^ The category gene limits were set at a minimum of 3 and a maximum of 2,000 genes. The analysis included 1,000 permutations, and from this, we interpreted the top 10 most significant biological processes and cellular components. Only terms that survived the false discovery rate-corrected statistical threshold of *P* < 0.05 were considered significant. To ascertain that the enrichments were specific to the eGFR-cortical thickness association, we repeated the same steps using instead body mass index, Framingham risk score or white matter lesion volume.

### 2.6. Spatial mapping analyses

We then performed spatial mapping analyses to investigate whether the association pattern between eGFR and cortical thickness mapped onto neurodegeneration.^38^ To do this, we used the previously published FreeSurfer-processed maps of changes in cortical thickness reported in Alzheimer’s disease and isolated REM sleep behavior disorder,^39^ a sleep disorder associated with abnormal and often violent movements while dreaming,^40^ where over 90% of patients develop a synucleinopathy such as Parkinson’s disease.^41, 42^ The maps were selected due to a similar T1-weighted MRI processing having been performed and a similar parcellation having been applied as in the present study.^39^ The map of Alzheimer’s disease comes from 78 patients who were part of the Alzheimer’s Disease Neuroimaging Initiative,^43, 44^ whereas the map of isolated REM sleep behavior disorder comes from 138 patients whose diagnosis was done based on published guidelines, including the use of sleep clinic-based polysomnography assessment.^45^ Spearman’s correlations were performed between the neurodegenerative brain maps and the t-value map of the association between eGFR and cortical thickness, independently from age, sex, body mass index, Framingham risk score, and white matter lesion volume.

### 2.7. Statistical analysis

#### 2.7.1. Demographic and clinical variables

The statistical analyses of demographic, clinical, and regional brain values were conducted using IBM SPSS Statistics (version 29.0). Baseline characteristics of the entire MRI cohort, as well as participants categorized into eGFR groups (see below), are presented as means and standard deviations for continuous variables and as proportions and percentages for categorical variables. To assess the presence of statistical differences between eGFR groups for continuous and categorical variables, between-group t-tests and chi-square tests were used, respectively.

#### 2.7.2. Association between eGFR and cognitive performance

Using the baseline CARTaGENE cohort with available eGFR and cognitive data, we conducted multivariate linear regressions to investigate the associations between eGFR and cognitive performance. The performance on the reaction time, working memory, and reasoning tasks was divided into quartiles, and regression models were used to determine the presence of significant differences in eGFR among quartiles. Similarly, participants were stratified based on baseline eGFR levels, and multivariate regression models were then applied to evaluate whether cognitive performance on the three tasks significantly varied across eGFR levels. These models were adjusted for age, sex, income, education, smoking, alcohol intake, hypertension, dyslipidemia, body mass index, diabetes, history of cardiovascular disease (including stroke), and psychoactive medication use.

#### 2.7.3. Cortical thickness analyses

Regression analyses of eGFR on cortical thickness at each vertex of the cortical surface were performed using general linear models, while controlling for age, sex, body mass index, Framingham risk score, and white matter lesion volume. To examine sex effects, we conducted vertex-based analyses to investigate the presence of differences between men and women in slopes while adjusting for the same covariates but sex.

Furthermore, to investigate the mediating effect of eGFR on the relationship between cognitive performance and cortical morphology, we looked for significant vertex-wise patterns linking cortical morphology and reaction time, working memory, and reasoning tasks, while adjusting for same covariates. We then conducted vertex-wise analyses to assess if regression slopes between cognitive performance and brain structure differed based on eGFR categories, defined as normal eGFR (≥90 ml/min/1.73 m^2^), mildly decreased eGFR (60-89 ml/min/1.73 m^2^), and moderately decreased eGFR (30-59 ml/min/1.73 m^2^). All vertex-wise surface analyses were performed separately for each hemisphere, and clusters were considered significant when *P* < 0.05 using a Monte Carlo simulation approach.

### 2.8. Patient Consents

All procedures were conducted in accordance with the ethical guidelines set forth in the Declaration of Helsinki and approved by institutional ethics committees of the Sainte-Justine University Health Centre and Hôpital du Sacré-Cœur de Montréal. Informed consent was obtained from all participants.

### 2.9. Data Availability

Qualified researchers may obtain access to all de-identified imaging data in the biomarkers registry used for this study (https://cartagene.qc.ca).

## 3. RESULTS

### 3.1. Demographics

A total of 19,996 participants were recruited in the CARTaGENE baseline cohort. Among these, 4,107 participants (20.5%) were excluded due to missing eGFR values or cognitive testing data or because they received cholinesterase inhibitors, yielding 15,897 participants to examine the relationship between eGFR and cognitive performance. A total of 1,397 participants underwent T1-weighted brain MRI 6 years after baseline. Of these, 60 (4.3%) were excluded due to missing clinical information or quality control, resulting in a sample of 1,336 participants.

On average, participants who underwent imaging were 53.9 ± 7.5 years, with 49% being women (Table 1). The time interval between baseline assessment and MRI acquisition was 6.2 ± 0.9 years. A total of 634 participants (47.5%) had normal eGFR (G1), 659 (49.3%) had mildly decreased eGFR (G2), and 43 (3.2%) had moderately decreased eGFR (G3). The G3 group was older and exhibited higher body mass index, Framingham risk scores, and white matter lesion volumes compared to the G2 group. The G2 group also showed differences on these variables compared to the G1 group.

**Table 1.**
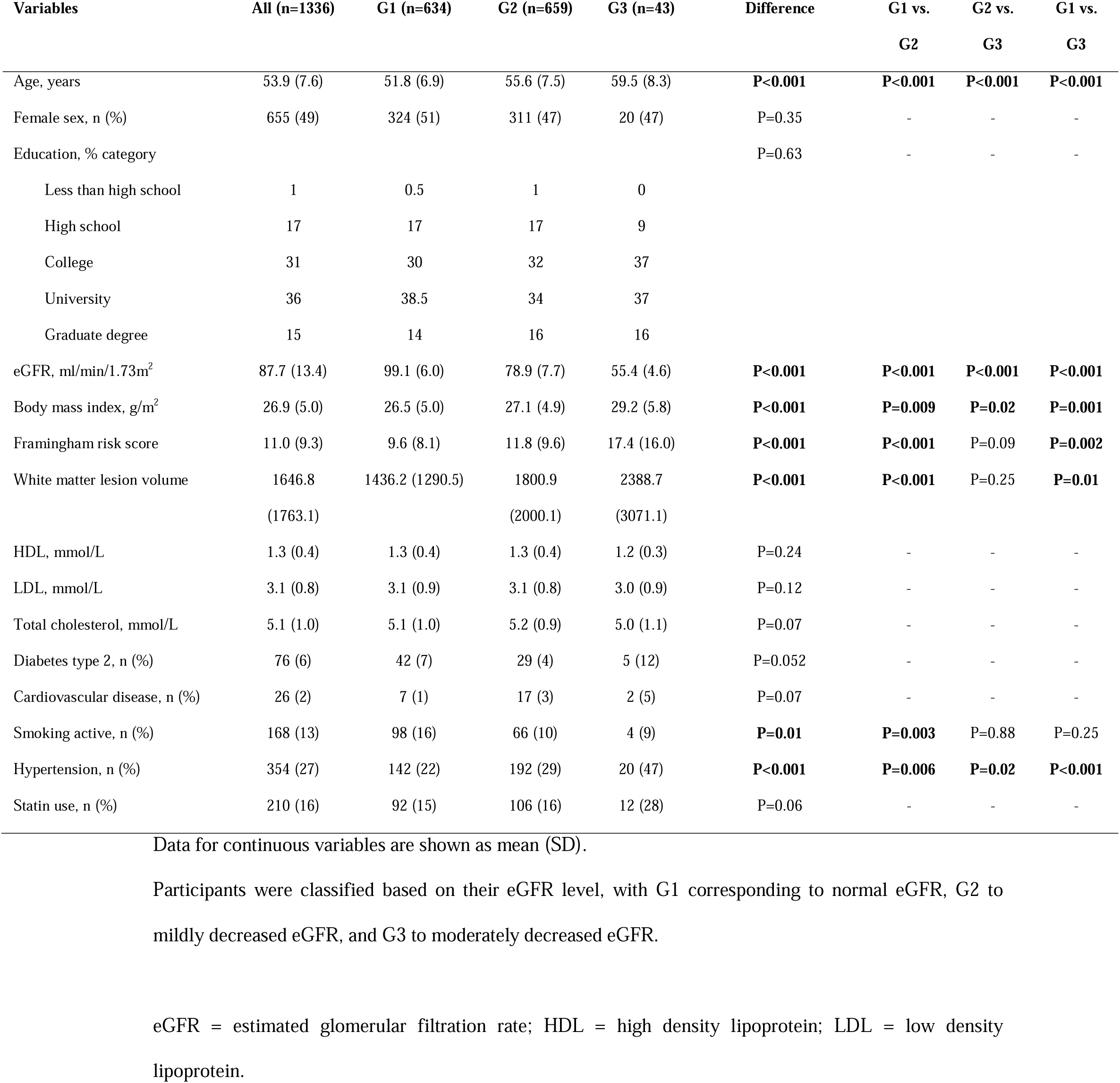
Demographics and clinical variables.

| Variables | All (n=1336) | G1 (n=634) | G2 (n=659) | G3 (n=43) | Difference | G1 vs.<br>G2 | G2 vs.<br>G3 | G1 vs.<br>G3 |
| --- | --- | --- | --- | --- | --- | --- | --- | --- |
| Age, years | 53.9 (7.6) | 51.8 (6.9) | 55.6 (7.5) | 59.5 (8.3) | **P<0.001** | **P<0.001** | **P<0.001** | **P<0.001** |
| Female sex, n (%) | 655 (49) | 324 (51) | 311 (47) | 20 (47) | P=0.35 | - | - | - |
| Education, % category |  |  |  |  | P=0.63 | - | - | - |
| Less than high school | 1 | 0.5 | 1 | 0 |  |  |  |  |
| High school | 17 | 17 | 17 | 9 |  |  |  |  |
| College | 31 | 30 | 32 | 37 |  |  |  |  |
| University | 36 | 38.5 | 34 | 37 |  |  |  |  |
| Graduate degree | 15 | 14 | 16 | 16 |  |  |  |  |
| eGFR, ml/min/1.73m<sup>2</sup> | 87.7 (13.4) | 99.1 (6.0) | 78.9 (7.7) | 55.4 (4.6) | **P<0.001** | **P<0.001** | **P<0.001** | **P<0.001** |
| Body mass index, g/m<sup>2</sup> | 26.9 (5.0) | 26.5 (5.0) | 27.1 (4.9) | 29.2 (5.8) | **P<0.001** | **P=0.009** | **P=0.02** | **P=0.001** |
| Framingham risk score | 11.0 (9.3) | 9.6 (8.1) | 11.8 (9.6) | 17.4 (16.0) | **P<0.001** | **P<0.001** | P=0.09 | **P=0.002** |
| White matter lesion volume | 1646.8<br>(1763.1) | 1436.2 (1290.5) | 1800.9<br>(2000.1) | 2388.7<br>(3071.1) | **P<0.001** | **P<0.001** | P=0.25 | **P=0.01** |
| HDL, mmol/L | 1.3 (0.4) | 1.3 (0.4) | 1.3 (0.4) | 1.2 (0.3) | P=0.24 | - | - | - |
| LDL, mmol/L | 3.1 (0.8) | 3.1 (0.9) | 3.1 (0.8) | 3.0 (0.9) | P=0.12 | - | - | - |
| Total cholesterol, mmol/L | 5.1 (1.0) | 5.1 (1.0) | 5.2 (0.9) | 5.0 (1.1) | P=0.07 | - | - | - |
| Diabetes type 2, n (%) | 76 (6) | 42 (7) | 29 (4) | 5 (12) | P=0.052 | - | - | - |
| Cardiovascular disease, n (%) | 26 (2) | 7 (1) | 17 (3) | 2 (5) | P=0.07 | - | - | - |
| Smoking active, n (%) | 168 (13) | 98 (16) | 66 (10) | 4 (9) | **P=0.01** | **P=0.003** | P=0.88 | P=0.25 |
| Hypertension, n (%) | 354 (27) | 142 (22) | 192 (29) | 20 (47) | **P<0.001** | **P=0.006** | **P=0.02** | **P<0.001** |
| Statin use, n (%) | 210 (16) | 92 (15) | 106 (16) | 12 (28) | P=0.06 | - | - | - |

Data for continuous variables are shown as mean (SD).
Participants were classified based on their eGFR level, with G1 corresponding to normal eGFR, G2 to mildly decreased eGFR, and G3 to moderately decreased eGFR.
eGFR = estimated glomerular filtration rate; HDL = high density lipoprotein; LDL = low density lipoprotein.

### 3.2. Baseline eGFR is associated with lower cognitive performance

In the 15,897 participants who underwent cognitive testing, we found a gradual decline on reaction time, working memory, and reasoning in relation to the eGFR levels (*P* < 0.001 for each task) after adjusting for age, sex, income, education, smoking, alcohol consumption, hypertension, dyslipidemia, body mass index, diabetes, a history of cardiovascular disease, and psychoactive medication usage (Table 2). When assessing the association inside each eGFR level separately, we observed a gradual decline in eGFR alongside lower cognitive performance on every task (reaction time: β*_eGFR_*= –6.6 (95% CI: –10.4 – –2.8), *P* = 0.001; working memory: β*_eGFR_* = –0.22 (95% CI: –0.4 – –0.027), *P* = 0.026; reasoning: β*_eGFR_* = –0.086 (95% CI: –0.035 – –0.014), *P* = 0.001).

**Table 2.** Associations between eGFR and cognitive performance.

| Cognitive task | Quartiles | Performance | Adjusted eGFR (SD),<br>ml/min/1.73 m<sup>2</sup> | Adjusted P-value |
| --- | --- | --- | --- | --- |
| Reaction time | Q1 | <0.43 s | 90.4 (5.2) | **P<0.001** |
|  | Q2 | 0.43-0.49 s | 88.5 (5.6) |  |
|  | Q3 | 0.49-0.57 s | 87.0 (6.0) |  |
|  | Q4 | >0.57 s | 85.6 (6.1) |  |
| Working memory | Q1 | 0-5 | 89.2 (5.7) | **P<0.001** |
|  | Q2 | 6-9 | 88.2 (5.9) |  |
|  | Q3 | 10-14 | 87.3 (6.0) |  |
|  | Q4 | >14 | 86.3 (6.1) |  |
| Reasoning | Q1 | ≥5 | 88.4 (5.7) | **P<0.001** |
|  | Q2 | 4 | 88.2 (5.8) |  |
|  | Q3 | 3 | 88.0 (5.9) |  |
|  | Q4 | ≤2 | 87.5 (6.2) |  |

Regression models were adjusted for age, sex, income, education, smoking, alcohol intake, hypertension, dyslipidemia, body mass index, diabetes, history of cardiovascular disease (including stroke), and psychoactive medication use. The first quartile (Q1) corresponded to the best cognitive performance, whereas the last quartile (Q4) related to the lowest cognitive performance. Bold values indicate a significant effect of eGFR.
eGFR = estimated glomerular filtration rate; Q = quartile; SD = standard deviation.

### 3.3. Baseline eGFR is associated with changes in brain morphology

Lower eGFR was associated with cortical thinning in frontal and posterior brain regions and thickening in temporal and cingulate areas (Fig. 1A). The relationship between lower eGFR and cortical thinning was significant in the left frontoparietal cortex, precuneus and cuneus, and in the right dorsolateral prefrontal cortex, precuneus, and inferior parietal cortex (Table S1 and Fig. 1A). There was no significant sex interaction.

**Figure 1.**
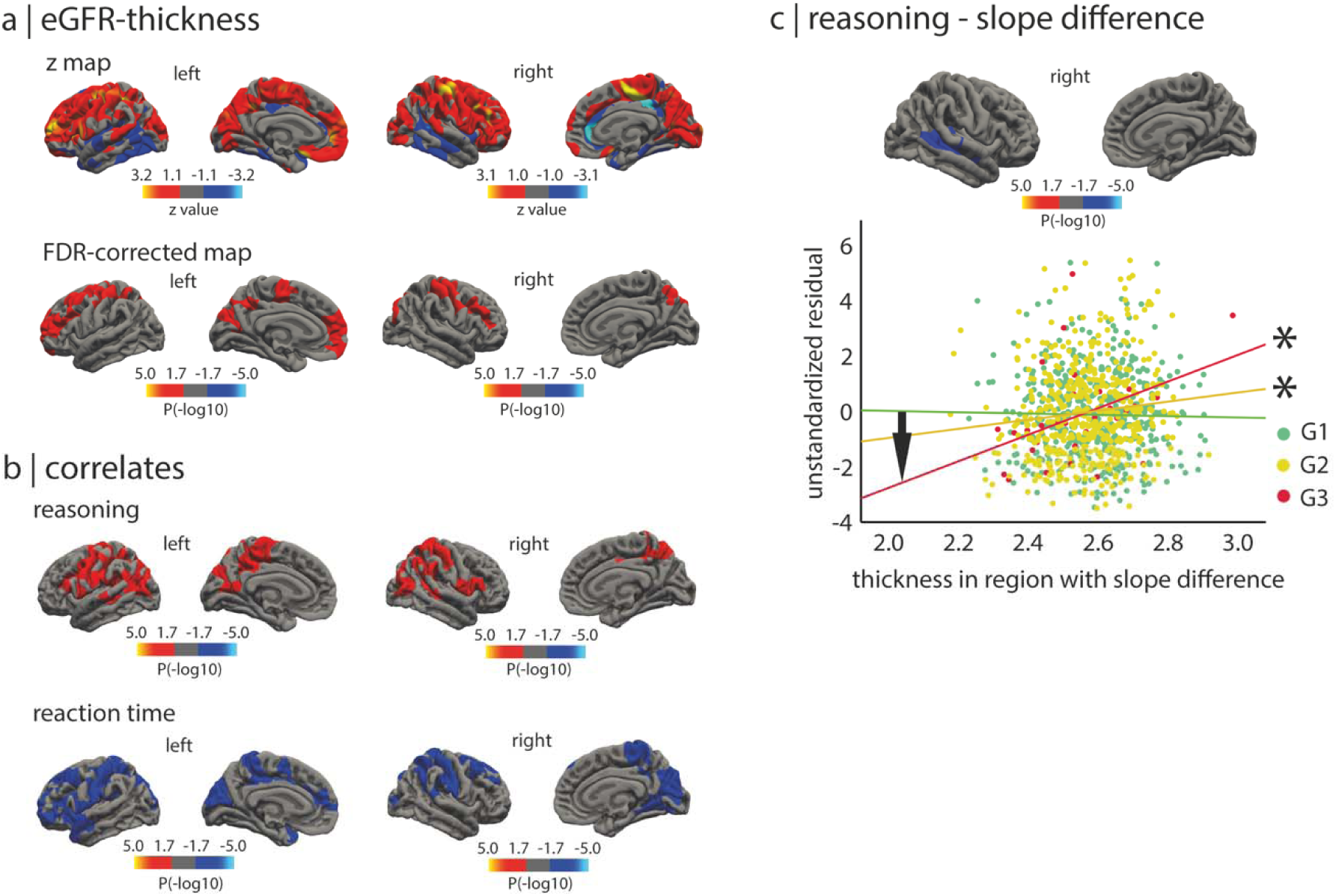
Associations between eGFR, brain structure, and cognitive performance. The eGFR is significantly associated with brain structure. **(A)** Vertex-wise analyses of eGFR against cortical thickness, with red areas representing the association between lower eGFR and thinning and blue areas the association with thickening. The uncorrected z maps and corrected FDR maps are shown. **(B)** Vertex-wise analyses of eGFR against performance on the reasoning and reaction time tasks, with red areas representing positive relationships and blue areas negative relationships. **(C)** Vertex-wise analyses showing where the association between reasoning performance and cortical thickness differed as a function of eGFR levels. The scatterplot shows the association between residualized reasoning performance and average thickness in the significant cluster. The asterisks represent the significant correlations, and the arrow indicates that the difference in the slopes between the G1 and G3 groups was significant. eGFR = estimated glomerular filtration rate; FDR = false discovery rate.

### 3.4. The eGFR mediates the relationship between cognition and morphology

Lower reasoning performance was correlated with cortical thinning in the bilateral sensorimotor, parietal, and posterior temporal cortices (Table S1 and Fig. 1B). Longer reaction times were associated with thinning in the bilateral prefrontal, anterior temporal, parietal, and occipital cortices (Table S1 and Fig. 1B). There were no associations between working memory and cortical thickness. The association between reasoning and cortical thickness was mediated by eGFR levels in the superior temporal cortex (Table S1 and Fig. 1C). This difference was significant between participants with normal eGFR and those with moderately decreased eGFR (Table S1 and Fig. 1C). While there was no association in individuals with normal eGFR (*r* = –0.01, *P* = 0.41), an association was present in those with mildly decreased eGFR (*r* = 0.11, *P* = 0.006) and more pronounced in those with moderately decreased eGFR (*r* = 0.46, *P* = 0.008) (Fig. 1C). The eGFR did not mediate the associations between thickness and working memory or reaction time.

### 3.5. Regional gene expression underlies eGFR-thickness associations

Lower eGFR correlated with thinning in the left medial orbitofrontal cortex (*r* = 0.08, *P_FDR_* = 0.0015) and thickening in the right rostral anterior cingulate cortex (*r* = –0.11, *P_FDR_* = 0.0007) (Table S2). Of the 5 gene expression components tested in our imaging transcriptomics pipeline (Fig. 2A), two components were significant, explaining respectively 36.8% (compared to 17.9% for null models, *P* = 0.006) and 23.2% (compared to 13.1% for null models, *P* = 0.048) of the variance between eGFR and thickness (Fig. 2B).

**Figure 2.**
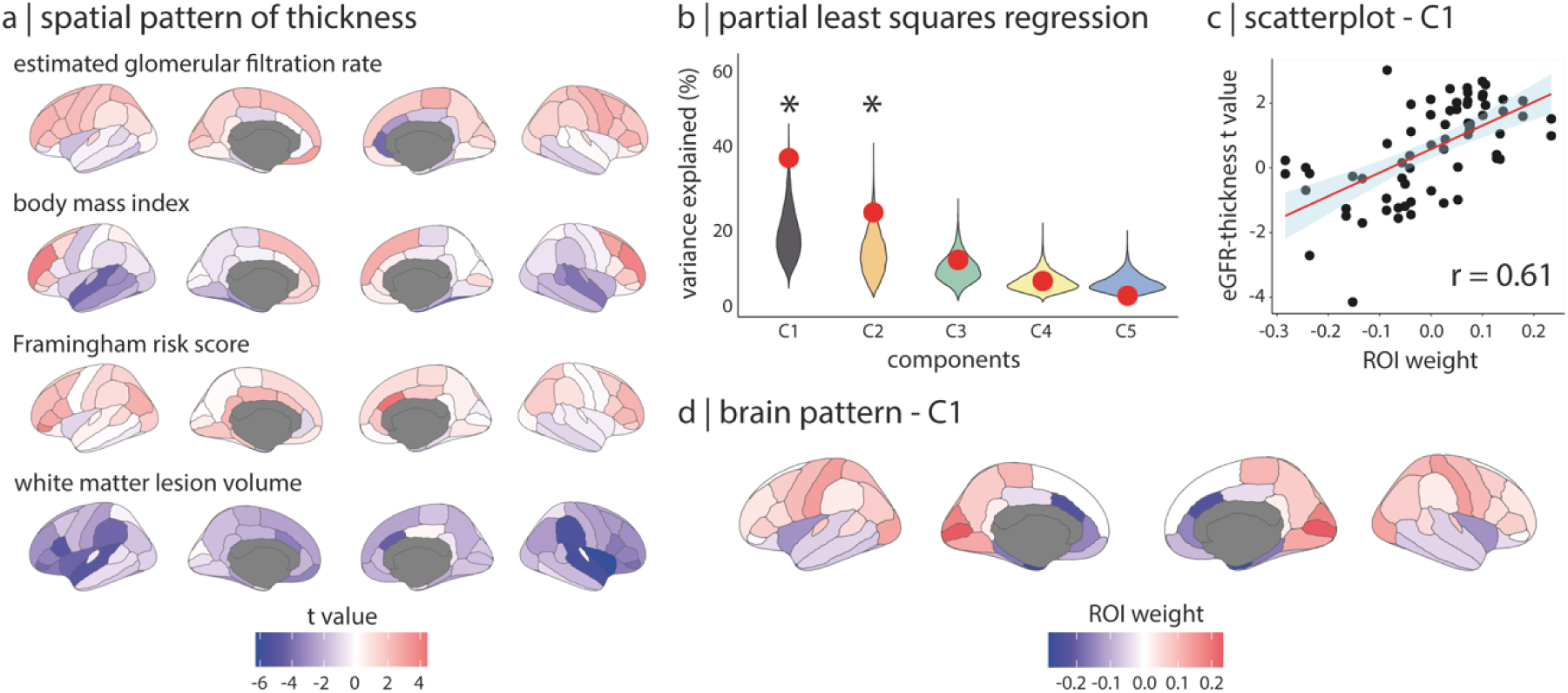
Regional gene expression patterns of the eGFR-thickness association. Regional gene expression patterns underlie the association between eGFR and cortical thickness. **(A)** Brain maps showing the spatial patterns of regional t-values between thickness and eGFR, body mass index, Framingham risk score, and white matter lesion volume independently from the effects of age, sex, and respective confounders. A positive t value represented the effect of decreased eGFR on thinning and a negative t value for the other variables represented the effect of an increase on thinning. **(B)** Two components explained significantly more variance in the eGFR-thickness spatial pattern than 10,000 spatial null models. **(C)** Scatterplot of the first component showing that regions where decreased eGFR associated with thinning (positive t-values) are positively associated with the component. **(D)** Brain map of the first component’s regional weights showing that regions positively associated with the component locate in the frontal, parietal, and occipital areas. C = component; eGFR = estimated glomerular filtration rate; ROI = region of interest.

For the first component, regions where lower eGFR correlated with thinning were positively weighted and found in the dorsolateral prefrontal and parietal cortices, while regions where lower eGFR correlated with thickening were negatively weighted and found in the temporal and anterior cingulate cortices (*r* = 0.61, *P* < 0.001, Fig. 2C, Fig. 2D). Regions where lower eGFR associated with cortical thinning had higher mitochondrial gene expression and overexpressed genes involved in mitochondrial RNA metabolic processes and potassium ion transport (Table 3 and Fig. 3A). Regions where lower eGFR associated with cortical thickening were enriched for genes involved in processes related to protein-containing complex remodeling (Table 3 and Fig. 3A), including apolipoprotein E and angiotensinogen. Enriched cellular components in regions with thinning involved mitochondrial and membrane elements, whereas glial cell projections and major histocompatibility complex (MHC) proteins were enriched in regions with thickening (Table S3). The second component linking eGFR and cortical thickness exhibited the same directional pattern as the first (*r* = 0.48, *P* < 0.001, Fig. 3B and 3C) but showed no enriched biological processes (Table S4). Specificity analyses revealed that gene enrichment patterns associated with body mass index, Framingham risk score, and white matter lesion volume were distinct from those found for eGFR (Fig. 2A), except for the glial cell projections also enriched for white matter lesion volume.

**Figure 3.**
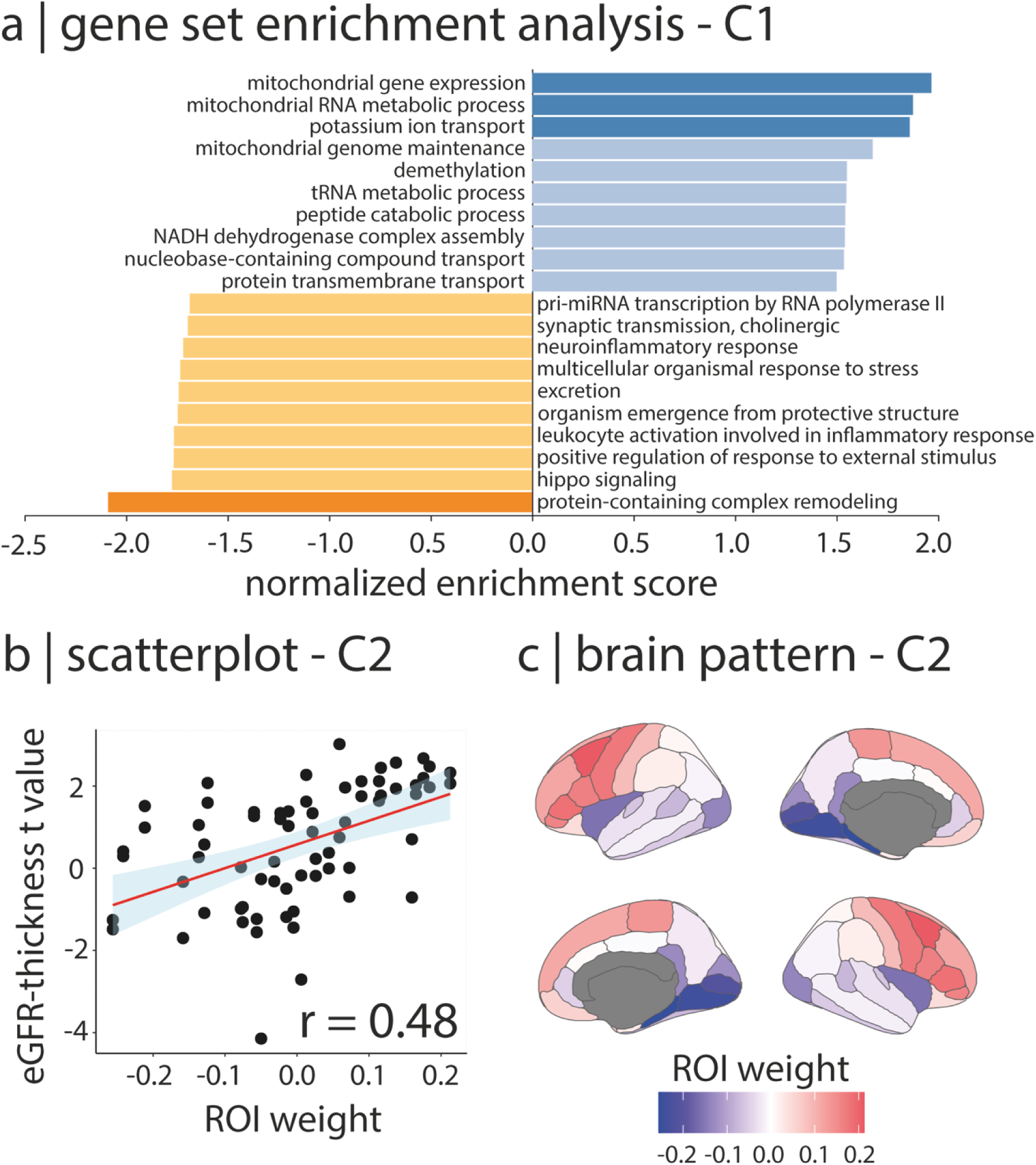
Gene set enrichment analysis of eGFR-thickness associations. Specific gene enrichments underlie the eGFR-thickness association in the brain. **(A)** Gene set enrichment analysis on the first component reveals biological processes enriched in regions where lower eGFR associated with thinning (blue) or thickening (yellow). Darker bars represent significance after FDR correction. **(B)** Scatterplot of the second component showing that regions where lower eGFR is associated with thinning (positive t-values) are positively correlated with the component. **(C)** Brain map of the second component’s regional weights showing that regions where lower eGFR associates with thinning (red areas) locate in the frontal and parietal regions. C = component; eGFR = estimated glomerular filtration rate; FDR = false discovery rate; pri-miRNA = primary microRNA; ROI = region of interest; RNA = ribonucleic acid.

**Table 3.** Biological processes enriched in the genes predicting the regional associations between eGFR and thickness.

| Gene set | Description | Size | Leading<br>edge<br>number | Enrichment<br>score | Normalized<br>enrichment<br>score | FDR *P*<br>value |
| --- | --- | --- | --- | --- | --- | --- |
| **Component 1: Positively weighted genes (more expressed with decreased cortical thickness)** |  |  |  |  |  |  |
| GO:0140053 | **mitochondrial gene expression** | 158 | 75 | 0.445 | 1.968 | **0.026** |
| GO:0000959 | **mitochondrial RNA metabolic process** | 40 | 22 | 0.539 | 1.877 | **0.041** |
| GO:0006813 | **potassium ion transport** | 188 | 62 | 0.406 | 1.861 | **0.035** |
| GO:0000002 | mitochondrial genome maintenance | 29 | 14 | 0.507 | 1.678 | 0.256 |
| GO:0070988 | demethylation | 49 | 21 | 0.418 | 1.551 | 0.347 |
| GO:0006399 | transfer RNA metabolic process | 173 | 91 | 0.342 | 1.549 | 0.323 |
| GO:0043171 | peptide catabolic process | 22 | 11 | 0.507 | 1.543 | 0.315 |
| GO:0010257 | NADH dehydrogenase complex assembly | 58 | 31 | 0.407 | 1.541 | 0.295 |
| GO:0015931 | nucleobase-containing compound transport | 217 | 70 | 0.332 | 1.536 | 0.286 |
| GO:1902600 | proton transmembrane transport | 121 | 34 | 0.347 | 1.500 | 0.349 |
| **Component 1: Negatively weighted genes (more expressed with increased cortical thickness)** |  |  |  |  |  |  |
| GO:0034367 | **protein-containing complex remodeling** | 17 | 6 | -0.673 | -2.089 | **0.025** |
| GO:0035329 | hippo signaling | 32 | 10 | -0.485 | -1.774 | 0.117 |
| GO:0032103 | positive regulation of response to external stimulus | 204 | 63 | -0.337 | -1.766 | 0.114 |
| GO:0002269 | leukocyte activation involved in inflammatory response | 22 | 8 | -0.531 | -1.765 | 0.103 |
| GO:0071684 | organism emergence from protective structure | 20 | 5 | -0.528 | -1.746 | 0.107 |
| GO:0007588 | excretion | 48 | 17 | -0.440 | -1.741 | 0.103 |
| GO:0033555 | multicellular organismal response to stress | 62 | 22 | -0.408 | -1.733 | 0.102 |
| GO:0150076 | neuroinflammatory response | 32 | 12 | -0.469 | -1.718 | 0.108 |
| GO:0007271 | synaptic transmission, cholinergic | 18 | 6 | -0.536 | -1.697 | 0.119 |
| GO:0061614 | pri-microRNA transcription by RNA polymerase II | 36 | 14 | -0.456 | -1.687 | 0.121 |

The top 10 biological process terms associated with the genes enriched in the association between eGFR and cortical thickness independently from the effects of age, sex, body mass index, Framingham risk score, and white matter lesion volume are presented ranked based on the normalized enrichment score. The terms showing significant enrichment after FDR correction are indicated in bold.
eGFR = estimated glomerular filtration rate; FDR = false discovery rate; NADH = nicotinamide adenine dinucleotide (reduced form); RNA = ribonucleic acid.

### 3.6. The associations map over regions associated with neurodegeneration

Using atrophy patterns of Alzheimer’s disease (66.0 ± 4.9 years, 51% women) and isolated REM sleep behavior disorder (67.0 ± 6.3, 19% women),^39^ an early pathognomonic feature of dementia with Lewy bodies and Parkinson’s disease,^41^ there was a near-significant spatial correlation between the brain map of regions where lower eGFR related to increased thickness and the atrophic regions in Alzheimer’s disease (*r* = 0.23, P = 0.056). There was a significant correlation between the brain map of regions where lower eGFR related to decreased thickness and atrophic regions in isolated REM sleep behavior disorder (*r* = –0.48, P < 0.001).

## 4. DISCUSSION

In this study, lower eGFR was associated with lower cognitive performance on reasoning, working memory, and reaction time tasks. These relationships were associated with structural brain changes, with eGFR mediating the relationship between reasoning and brain changes. Our imaging transcriptomic analysis revealed that lower eGFR correlated with specific transcriptomic patterns, including the atrophy patterns seen in tauopathies and synucleinopathies.

CKD has been associated with cognitive decline, an increased risk of developing neurodegenerative conditions, and changes in brain structure.^3, 4, 9^ In this study, we confirmed, in a large population-based cohort representative of the general population, the previous observations that lower eGFR is associated with reduced cognitive performance, specifically in reasoning and memory tasks, along with longer reaction times. This is consistent with prior research indicating that CKD impacts various cognitive domains, with a notable effect on attention and executive functions.^4^ Furthermore, we observed that the association of lower eGFR with reduced cognitive performance correlated with brain structure, specifically in the reasoning task and only in individuals with decreased eGFR levels, with a worsening effect on cognition as eGFR decreased. Importantly, in contrast to previous studies that investigated eGFR by including individuals with diagnosed CKD, our findings were obtained in individuals who still have healthy levels of eGFR, meaning that our observations represent very early associations between kidney function, cognitive performance, and brain structure.

We found that brain areas where lower eGFR related to increased cortical thickness displayed an enrichment of genes associated with the protein-containing remodeling complex and tended to map on the atrophy associated with Alzheimer’s disease. The *APOE* gene, which is part of the protein-containing remodeling complex, was the gene most robustly associated with the lower eGFR-increased thickness relationship. The *APOE* ε4 variant is one of the greatest risk factors for the development of Alzheimer’s disease.^46^ This gene was followed by *AGT*, which codes for angiotensinogen, the precursor to active forms of angiotensin involved in regulating blood pressure, fluid balance, and neuroinflammation.^47^ In line with that, regions where increased cortical thickness associated with lower eGFR were enriched for the major histocompatibility complex, a marker of microglia activation. Proteins levels of molecules from this complex are increased in Alzheimer’s disease brains and inversely correlated with cognitive function. ^48^ Importantly, we found that the association between lower eGFR and brain changes were distinct from those linking eGFR with vascular risk factors, white matter lesion volume, and body mass index. This supports that the increased cortical thickness may be a signal of the effect of inflammation in regions that align with tauopathies.

Regions where lower eGFR related to cortical thinning presented with an enrichment of mitochondrial gene expression. These regions also mapped with the atrophy seen in prodromal synucleinopathies, namely isolated REM sleep behavior disorder. Isolated REM sleep behavior disorder is a prodromal phenotype increasingly recognized as a harbinger of Parkinson’s disease and dementia with Lewy bodies,^49^ with over 90% of patients developing either one of these diseases within 15 years.^41^ The atrophy associated with this phenotype occurs in regions with a higher expression of genes involved in oxidative phosphorylation,^39^ which are akin to the mitochondrial gene expression and RNA metabolic processes identified in relation to lower eGFR in this study. While not indicative of causal pathways between lower eGFR and synucleinopathies, our findings are in line with the notion that mitochondrial involvement represents a shared commonality in the pathophysiology associated with synucleinopathies and the kidney-brain axis. Indeed, previous studies reported an association between lower eGFR and higher probability of prodromal Parkinson’s disease and parkinsonism.^5, 7^ A possible explanation for the shared pathways between lower eGFR and neurodegenerative conditions could be the common influence of increased inflammation and oxidative stress on the brain.^8, 50^ In CKD, alterations in the renin-angiotensin system contribute to neurodegeneration by raising circulating angiotensin II levels, increasing blood pressure, and potentially compromising the blood-brain barrier.^8^ The binding of angiotensin II to the brain’s angiotensin receptors, as supported by our demonstration of an enrichment of the gene *AGT* in the regions showing a lower eGFR-increased thickness relationship, may trigger local oxidative stress and microglial activation,^8^ possibly leading to neuronal damage and structural changes in the brain.

This study has some limitations. First, brain atrophy relied on T1-weighted imaging, limiting our understanding of white matter tracts. Future investigations should consider incorporating diffusion MRI and advanced probabilistic tractography to explore the intricate relationships between eGFR, cognitive function, and the impact of white matter changes on gray matter alterations. Second, the calculation of eGFR was based solely on creatinine levels. However, previous research that measured eGFR through creatinine, cystatin C, combined creatinine-cystatin C or β_2_-microglobulin demonstrated that these do not significantly alter the interpretation of eGFR in relation to brain measurements.^12^ Third, we adjusted for cardiovascular risk factors using the composite Framingham risk score, a decision made to mitigate the potential loss of statistical power associated with introducing categorical variables for individual risk factors such as diabetes, hypertension, and dyslipidemia. This approach allowed considering vascular risk factors together, without modeling class-dependent parameters in the general linear models. Fourth, most participants had an eGFR within a relatively narrow range and had very mild CKD, suggesting that our results may not be generalizable to persons with more

severe kidney disease. Nonetheless, even within this narrow range of mostly mild CKD, a significant association was seen between eGFR, cognitive test results, and brain imaging.

In conclusion, this study demonstrates the relationship between eGFR, cognitive performance, and brain structure in healthy individuals from a large population-based cohort. It supports the idea that the effect of eGFR on brain structure is not random and targets regions with distinct gene expression patterns.

## 5. ACKNOWLEDGMENTS

The authors want to thank all participants included in the CARTaGENE cohort. R.G. holds a research scholarship from the Fonds de recherche du Québec – Santé and is a recipient of the Société québécoise d’hypertension artérielle – Bourse Jacques-de-Champlain scholarship.

## Notes

### Competing Interest Statement

The authors have declared no competing interest.

### Funding Statement

This study did not receive any funding

### Author Declarations

IRB of the McGill University Health Centre and the Centre integre universitaire de sante et de services sociaux du Nord de l'Ile de Montreal

